# Trajectory of severe COVID anxiety and predictors for recovery in an 18-month cohort

**DOI:** 10.1101/2024.07.22.24310664

**Authors:** Jacob D King, Aisling McQuaid, Kirsten Barnicot, Paul Bassett, Verity C Leeson, Martina Di Simplicio, Peter Tyrer, Helen Tyrer, Richard G Watt, Mike J Crawford

**Author notes:** **Corresponding author:** Dr Jacob D King. Division of Psychiatry, Imperial College London. Floor 2, Commonwealth Building, Hammersmith Hospital, Du Cane Road, London W12 0HS. **Declaration of Interest:** None. **Authors’ contributions:** All authors meet ICJME criteria for authorship. MJC, VCL, KB, MDS, PT, HT, and RW designed the COVID Anxiety Project. AM and JDK collected data and VCL coordinated the study. PB and JDK conducted data analysis, and had full access to raw data. JDK drafted initial versions of this manuscript. All authors reviewed and approved a final version of this manuscript. **Transparency declaration:** Authors affirm that this manuscript is an honest, accurate and transparent account of the study as planned. **Availability of data and materials:** Data relating to the present analysis are available from the corresponding author on reasonable request. In due course, on completion of analyses, the full COVID Anxiety Project dataset will be made publicly accessible. **Ethics approval and consent to participate:** The study received approval from the Leicester Central Research Ethics Committee and Health Regulation Authority in 2020, reference number 20/EM/023. Research was conducted in line with the Declaration of Helsinki 1964, and responses were managed following General Data Protection Regulations. All individuals were required to provide informed consent, by independently reading a participant information sheet before signing an online consent form to participate. Participants received a copy of their consent form.

## Abstract

**Background:** People with severe COVID anxiety have significant fears of contagion, physiological symptoms of anxiety in response to a COVID stimuli, and employ safety behaviours which are often in excess of health guidelines and at the expense of other life priorities. The natural course of severe COVID anxiety is not known.

**Methods:** This prospective cohort study followed 285 people with severe COVID anxiety in United Kingdom over 18-months. Descriptive statistics and linear regression models identified factors associated with change in COVID anxiety.

**Results:** Most participants experienced major reductions in COVID anxiety over time (69.8% relative cohort mean decrease; p<0.001), but a quarter of people (23.7%, 95% CI 17.8 to 30.1) continued to worry about COVID every day. Increasing age, being from an ethnic background which conferred greater risk from COVID-19, and the persistence of high levels of health anxiety and depressive symptoms predicted significantly slower improvements in severe COVID anxiety adjusting for other clinical and demographic factors.

**Conclusions:** For most people severe COVID anxiety significantly improves with time. However established interventions treating depression or health anxiety, and targeting older people and people from at-risk minority groups who appear to recover at slower rates, might be clinically indicated in future pandemics.

**Highlights:**

- Most people with severe COVID anxiety reported large improvements in symptoms 18-months later.
- Levels of co-occurring poor mental health and social functioning also improved for most people.
- More than 1 in 10 continued to have severe COVID anxiety symptoms 18 months later.
- Age, ethnic background, and high levels of health anxiety and depression predict slower improvements.
- People with these risk characteristics could be considered for targeted support.

## Background

Anxious thoughts and feelings about COVID-19 are common, and appropriate given the very real threats to health and wellbeing posed by the virus. But for around 5-15% of people at the height of the pandemic, fears of contagion and anticipation of adverse physical and socioeconomic consequences became overwhelming, leading to changes in behaviour which were no longer adaptive, in ways that reinforced preoccupation with contagion. (1, 2) These included excessive consumption of COVID related news media, online symptom checking, hand washing, disinfecting items brought into the home and not leaving the home to a degree disproportionate to public health guidance, which stopped people from being able to get on with other life priorities. (3, 4, 5) Moreover somatic symptoms of anxiety like shortness of breath and gastrointestinal disturbance were sometimes misinterpreted as signs of COVID-19 itself, further reinforcing bodily vigilance, and symptom-checking behaviours which perpetuated anxiety.

Greater levels of COVID anxiety have been associated with poorer levels of overall mental health, socio-occupational functioning, and a lower quality of life, as well a greater rates of generalised, death, and health anxiety, post-traumatic stress, obsessive-compulsive, and depressive symptoms (3, 6), and substance misuse. (7, 8) In this sense people with the most severe degree of COVID anxiety may benefit from specific mental health support. COVID anxiety also appears to be an heterogenous experience, insofar as there have emerged different underlying psychological processes at play (2, 9), and specific clinical clusters associated with patterns of associated health behaviours and demographic profiles. (10)

There has now been much written on the changes in population general mental health, pre, during, and post the coronavirus pandemic, with near global consistence in patterns of significant increases in population levels of psychological distress and specific anxiety disorders which, having peaked in the early stages of the pandemic followed a gradual return to near pre-pandemic levels. (11, 12, 13) Specific groups of people appear to have had their rates poor mental health decline much more slowly, most notably adolescents and young people (14), and people who had existing poor mental health states prior to the pandemic. (15)

Yet only a handful of longitudinal studies which report the level of COVID anxiety (or closely related concepts like coronaphobia, COVID Stress Disorder/Syndrome, COVID-19 fear, COVID-related worries, or COVID-19 anxiety syndrome) in the same people over time have been published, and none have examined which factors are predictive of a change in symptoms. (16, 17, 18, 19) These studies ranged in their follow-up periods from between 5 and 14 months and cover a time period from February 2020 until July 2021 in China, Netherlands, United Kingdom and United States. Two studies assessed symptoms at more than 2 time-points. All studies demonstrated meaningful reductions in the level of COVID anxiety in time. However there do not appear to be studies which have assessed levels of COVID anxiety at a time after the coronavirus became endemic, when there were no longer public health restrictions in place and cases no longer routinely monitored. Moreover since the latest follow-up timepoint of these longitudinal studies there has been much greater uptake of vaccination, the development and availability of effective treatments and declining mortality rates, all factors likely to be associated with improving COVID anxiety. The natural course of post-pandemic severe COVID anxiety is unknown, as are the factors which might be associated with improvement or persistence of these symptoms over time.

## 2.0 Methods

The ‘COVID Anxiety Project’ recruited adults with severe COVID anxiety from United Kingdom during the British second and third waves between January and September 2021. This was a time more than a year after the first UK cases were identified, during which the third national lockdown had ended and social distancing restrictions eased, vaccination was increasingly available, but case rates remained high. Participants were followed up three, six and 18 months after recruitment, with the last follow-up completed in April 2023 when there was no longer national tracking of case rates and no public health restrictions in place. Participants received a £10 e-voucher for completion of the baseline survey and a £20 e-voucher for completion of the surveys at 6 and 18 month follow-ups.

### 2.1 Participants

1,068 individuals who self-identified as anxious about COVID responded to adverts on social media platforms and from contact made by 19 affiliated family medicine clinics in London. 306 people (30.4%) scored nine or more on the 20-point COVID Anxiety Scale (CAS)(20) indicating severe COVID anxiety, provided written consent and met other eligibility criteria including not having a history of psychotic illness, nor having had COVID-19 in the preceding four weeks. (21) The cohort recruited participants until there were enough people meeting further criteria to power a nested randomised feasibility trial of CBT for health anxiety. (22)

### 2.2 Measures

We assessed the level of COVID anxiety using the self-report 5-item CAS, which was the first, and at the time most widely used, validated assessment scale. This scale assesses the frequency of five somatic symptoms (loss of appetite, gastrointestinal disturbance, poor sleep, dizziness, and feeling paralysed or frozen) experienced in response to a COVID stimuli, like watching or reading news about the pandemic. A score of five or more indicates the likely presence of levels of COVID anxiety associated with significant disruption to daily life, and a score of nine or more a severe degree of symptomatology. (23) At baseline self-complete questionnaires were used to assess for symptoms of generalised anxiety, depression, health anxiety, obsessive-compulsive symptomatology, personality disorder, and drug and alcohol use. Respectively these were the Generalised Anxiety Disorder scale – 7 items (GAD-7) (24), Patient Health Questionnaire – 9 items (PHQ-9)(25), the 14 health anxiety items of the Short form Health Anxiety Inventory (sHAI) (26), Obsessive-Compulsive Inventory – Revised (OCI-R) (27), the Standardised Assessment of Personality Abbreviated Scale (SAPAS)(28), Drug use with the Single Drug Use Item (SDUI)(29), and the Alcohol Use Disorders Identification Test for Consumption (AUDIT-C). (30) There was good internal consistency of these measures at baseline assessed by Cronbach’s coefficient alpha (α > 0.8), aside for the SAPAS (α = 0.55).

Additionally we used measures of social and occupational functioning (Work and Social Adjustment Scale (31)), quality of life (EuroQuol 5-Domains (EQ-5D-3L) (32)) and novel measures of COVID specific health behaviours including hand washing, staying at home, disinfecting items brought in to the home, and consuming media relating to COVID-19 which were developed in association with people with lived experience of poor mental health during the pandemic. (3) Demographic factors were collected including age, sex, ethnicity, employment and relationship status, household composition, as well as self-disclosed physical health conditions which were coded using MedDRA criteria (https://www.meddra.org/) by a physician. Health conditions and participants’ ethnic background were assessed as increasing an individual’s risk to COVID-19 with the QCOVID tool (https://qcovid.org/). 21 participants were excluded from these analyses as they received a course of adapted CBT for Health Anxiety as part of the nested feasibility trial which we anticipated would influence the natural trajectory of severe COVID anxiety. (21)

There were significant rates of mental health conditions in this cohort, and otherwise nationally representative rates of at-risk health conditions. A full description of baseline cohort characteristics has been reported previously. (3) Latent profile analysis identified four clinically distinct clusters of people with severe COVID anxiety. (10) Clusters with significantly lower and higher burdens of co-occurring mental health symptoms emerged, along with a cluster of individuals with greater than expected rates of at-risk health conditions who were older than cohort average and abstinent of alcohol, and a fourth with prominent obsessive-compulsive symptoms and personality pathology described as ‘anankastic’.

At the 18-month follow-up point we collected repeat measures for COVID anxiety, generalised and health anxiety, depression, drug and alcohol use, and social-occupational dysfunction. All data was collected using the Qualtrics platform (https://www.qualtrics.com/en-gb/).

### 2.3 Data analysis

We published a statistical analysis plan in the ISRCTN registry prior to analysis (ISRCTN14973494). There was no within-scale missing data as the data collection platform that we used required that participants complete all items before they could progress through the survey.

We used descriptive statistics and linear regression models to assess the change in CAS scores from baseline to the 18-month follow-up. Univariate analysis first examined the unadjusted associations between change in CAS score and demographic factors, and clinical factors both as baseline predictors and the reductions in scores in the clinical measures between baseline and the 18-month time-point. We used a backwards selection procedure in multivariate analysis looking at the joint association of only those factors found to be associated with the outcome in univariate analysis (p<0.2). All data were managed and analysed in SPSS version 20.0 (SPSS Inc., Chicago IL) and Stata version 16.1 (College Station: StataCorp LLC; 2021).

### 2.4 Ethical approval

This study was reviewed favourably by the Leicester Central Research Ethics Committee and Health Regulation Authority in 2020, reference number 20/EM/023. We followed the Declaration of Helsinki 1964, and responses were managed following General Data Protection regulations. Upon recruitment all individuals were required to read and sign a consent form to participate, and received a copy.

## 3.0 Results

Of 285 participants, 177 (61.2%) provided responses at 3-month follow-up, 204 (71.2%) at 6 months and 199 (69.8%) at 18 months. Men (p=0.008), people of a Black, South Asian or Mixed ethnic heritage (p=0.03), people with baseline scores indicating lower levels of personality pathology (p=0.03) and those in the sub-group without high rates of co-occurring psychopathology (p=0.03) were less likely to have completed follow-up at 18 months.

Cohort mean average CAS scores dropped from 12.4 (95% CI 12.1 to 12.7), to 7.7 (95% CI 7.1 to 8.4) at 3 months, 6.8 (95% CI 6.2 to 7.4) at 6 months, and 3.7 (95% CI 3.2 to 4.2) at 18 months (a 69.8% relative decrease; p<0.001). 13.1% (n=26) of the cohort continued to score over 9 (indicating severe COVID anxiety) at 18 months, and 38.2% (n=76) continued to score 5 or more indicating levels of COVID anxiety associated with an impact on daily life. 61 (31%) people scored 0 at 18-months, indicating resolution of symptoms.

The cohort demonstrated major reductions in mean generalised anxiety (−31.7%, p<0.001), depressive symptoms (−24.5%, p<0.001), health anxiety (−23.9%, p<0.001), and social and occupational dysfunction (−43.0%, p<0.001) over 18 months.

At the 18-month follow-up, almost half of the sample reported still washing their hands a lot more than before the start of the pandemic (47.9%, 95% CI 40.6 to 55.3), 1 in 5 continued to disinfect most items coming into their home (21.6%, 95% CI 16.0% to 28.1%), another 1 in 5 reported watching COVID-related news media at least daily (22.1%, 95% CI 16.4% to 28.7%), and almost a quarter (23.7%, 95% CI 17.8 to 30.1) reported that they still worried about COVID every day. Of those with school-age children 10% continued not to send their children to school, even though they were expected to, because of concerns of COVID contagion.

When adjusted for baseline CAS score age was the only demographic factor which remained significant (p=0.005) at alpha level 0.05 in univariate analysis, as being predictive of slower improvements in COVID anxiety symptoms over time. While baseline levels of generalised anxiety, depression, personality pathology, obsessive-compulsive symptoms and alcohol use were also not predictive of a differing trajectory, improvements in generalised anxiety, health anxiety and depression over time were favourably and significantly associated with a CAS reduction. Other variables including living with an at-risk health condition or living with someone vulnerable to COVID-19, were not associated with smaller reductions in COVID anxiety over time. There was no predictive effect of cluster on magnitude of CAS reduction, nor was there an effect of vaccination status or having contracted COVID-19.

In multivariate modelling, age, and coming from a Black, South Asian or Mixed ethnic background were significant predictors of lesser reductions in CAS. Every 10-year increment increase in age was associated with a 0.4-point smaller reduction in CAS score over 18 months. Furthermore, individuals for whom depressive and health anxiety symptoms reduced more over the follow-up period also had significantly greater reductions in their COVID anxiety.

## 4.0 Discussion

This study has shown that among a self-selecting sample of people with severe COVID anxiety there were meaningful improvements in symptoms over time, and for around 1 in 3 a complete resolution. There were corresponding declines in co-occurring mental health symptoms and in the unhelpful forms of COVID protective behaviours. However, for almost 4 in 10 there remained a degree of COVID anxiety which had an impact on daily life, and more than 1 in 10 continued to experience a severe degree of symptoms 18-months later. This, at a time more than one year after the ending of any and all social restrictions and public health guidance in United Kingdom. Moreover despite significant improvements in co-occurring mental health scores over time, this population’s 18 month PHQ-9 and GAD-7 mean scores were more than double the nationally representative figures over this period (33). This might suggest that people who at one time experienced severe levels of COVID anxiety may also be a population with longer standing mental health difficulties, or who developed persistent mental health difficulties after their COVID anxiety improved.

This is the first study which has examined the trajectory and factors associated with changes in COVID anxiety over a long follow-up period among people with the most severe levels of COVID anxiety, and corroborates other studies’ findings that COVID related anxiety improves progressively in time. Previous studies have identified a host of factors which appear associated with the severity of COVID fear during the pandemic including older age, non-White ethnicity, personality traits, female gender, general level of both physical and mental health. (16, 18, 34, 35, 36) Our study builds on this by suggesting that increasing age and being from an at-risk ethnic background also appear to prolong the severity of COVID anxiety, but that other factors which contribute to individuals’ COVID-19 risk profile, including having an at-risk health condition and vaccination status, do not appear to affect the trajectory of severe COVID anxiety.

We previously conducted analysis on this same cohort at the 6-month time-point (22) which identified that older age, higher baseline levels of generalised and health anxiety, and greater reductions in health anxiety and depressive symptoms were significantly associated in multivariable models with greater reductions in COVID anxiety. These findings are again demonstrated one year later at the 18-month follow-up timepoint, however the effect of baseline mental health scores become no longer predictive of the longer-term trajectory of severe COVID anxiety. In this light, we suggest that older adults and people from minority ethnic backgrounds ought be held in special consideration for targeted support given that these factors appear associated with both increased severity of COVID anxiety, and its persistence.

Studies exploring the associations between COVID anxiety and symptoms of other mental health disorders appear to show a complex connection, likely bidirectional between COVID anxiety, and other anxiety states (37, 38), indeed a more general psychological distress factor may precede the expression of groupings of mental health symptom during the pandemic. Given our findings that greater decreases in depressive and health anxiety symptoms were associated with greater declines in COVID anxiety, we might suggest that there is cause to believe that established mental health interventions proven to reduce these symptoms have a theoretical basis for also improving COVID anxiety. And to note reciprocally, that randomised clinical trials of interventions specifically targeting COVID anxiety, particularly online CBT based interventions (22, 39, 40) appear to improve other mental health outcomes too.

## 5.0 Limitations

This study is limited in several ways. Firstly in that it was not nationally representative, and that our recruitment procedure, using mainly online and text message-based adverts and an on-line platform may have posed a barrier to certain groups, perhaps older adults and those without access to the internet, in hearing of the study and engaging with it. Second, we could not account for changes in participants’ mental health prior to the start of the pandemic nor the trajectory their mental health may have been on before recruitment. Third, that some factors likely to partially explain our findings went uncollected. For example we did not collect data on whether participants underwent mental health support during this period, nor their levels of social support and connectedness, which has appeared to play a role in predicting the severity of COVID anxiety in other studies. (41, 42, 43)

## 6.0 Conclusions

Almost 40% of people with debilitating thoughts and feelings about COVID-19 during the British third-wave of the pandemic continued to experience levels of COVID anxiety associated with social and occupational dysfunction 18 months later, and for 13% this remained severe. Older adults and people from ethnic backgrounds which conferred greater risk from COVID-19 were significantly slower to feel better, and ought be considered for targeted support. Reducing co-occurring health anxiety and depressive symptoms with established interventions might be a target for supporting adults with pandemic-related anxiety in the future.

## Supporting information

[Table 1 inserted here]

[Table 2 inserted here]

[Table 3 inserted here]

## Data Availability

Data relating to the present analysis are available from the corresponding author on reasonable request. In due course, on completion of analyses, the full COVID Anxiety Project dataset will be made publicly accessible.

## Acknowledgements

We are grateful to members of the Combined Independent Oversight Committee which oversaw project governance, data management and review safety data (Dr John Green (Chair), Professor Khalida Ismail, and Mr Robert Koch). We are grateful to MQ Research and Anxiety UK for helping to publicise the study. We also wish to thank members of the lived experience reference group including Manisha Ahya, Charlotte Green, Sandra Jayacodi, Niruben Patel and Vikas Sharma for their contributions to contextualising the results of this study and in the development of novel COVID-19 health behaviour items.

## Author details

**Dr Jacob D King**. Division of Psychiatry, Imperial College London.

**Ms Aisling McQuaid**. Division of Psychiatry, Imperial College London.

**Dr Kirsten Barnicot**. School of Health Sciences, City University of London.

**Mr Paul Bassett**. StatsConsultancy Ltd, Amersham, UK.

**Dr Verity C Leeson**. Division of Psychiatry, Imperial College London.

**Dr Martina Di Simplicio**. Division of Psychiatry, Imperial College London.

**Prof Peter Tyrer**. Division of Psychiatry, Imperial College London.

**Dr Helen Tyrer**. Division of Psychiatry, Imperial College London.

**Prof Richard G Watt**. Epidemiology and Public Health, University College London.

**Prof Mike J Crawford**. Division of Psychiatry, Imperial College London.

