## Supplementary material for "Trajectory of severe COVID anxiety and predictors for recovery in an 18-month cohort": [Table 1 inserted here]

| Variable | Timepoint | n | Mean ± SD | Change  Mean (95% CI) | P-value vs. baseline |
| --- | --- | --- | --- | --- | --- |
| CAS | Baseline | 285 | 12.4 ± 3.0 | 0 |  |
|  | 3 months | 177 | 7.7 ± 4.4 | -4.6 (-5.2, -4.1) | **<0.001** |
|  | 6 months | 204 | 6.8 ± 4.5 | -5.5 (-6.0, -4.9) | **<0.001** |
|  | 18 months | 199 | 3.7 ± 3.9 | -8.7 (-9.2, -8.1) | **<0.001** |
| WSAS | Baseline | 267 | 21.3 ± 7.7 | 0 |  |
|  | 3 months | 169 | 19.0 ± 9.0 | -2.7 (-3.9, -1.4) | **<0.001** |
|  | 6 months | 203 | 18.3 ± 9.0 | -3.3 (-4.5, -2.1) | **<0.001** |
|  | 18 months | 199 | 12.1 ± 9.5 | -9.4 (-10.6, -8.2) | **<0.001** |
| GAD-7 | Baseline | 274 | 15.4 ± 4.0 | 0 |  |
|  | 3 months | 172 | 13.4 ± 5.0 | -2.1 (-2.9, -1.4) | **<0.001** |
|  | 6 months | 203 | 12.5 ± 5.5 | -3.0 (-3.7, -2.3) | **<0.001** |
|  | 18 months | 199 | 10.5 ± 6.1 | -5.1 (-5.8, -4.4) | **<0.001** |
| sHAI | Baseline | 269 | 23.2 ± 7.1 | 0 |  |
|  | 3 months | 171 | 21.7 ± 8.5 | -1.8 (-2.6, -0.9) | **<0.001** |
|  | 6 months | 203 | 19.7 ± 8.4 | -3.5 (-4.3, -2.6) | **<0.001** |
|  | 18 months | 199 | 18.2 ± 8.1 | -5.0 (-5.8, -4.1) | **<0.001** |
| PHQ-9 | Baseline | 268 | 15.6 ± 5.5 | 0 |  |
|  | 3 months | 170 | 13.8 ± 6.4 | -1.5 (-2.3, -0.6) | **0.001** |
|  | 6 months | 203 | 13.4 ± 6.7 | -2.1 (-2.9, -1.3) | **<0.001** |
|  | 18 months | 199 | 11.7 ± 7.2 | -4.0 (-4.8, -3.1) | **<0.001** |
| OCI-R | Baseline | 264 | 30.4 ± 15.3 | 0 |  |
|  | 3 months | 169 | 28.2 ± 16.8 | -1.5 (-3.2, 0.2) | 0.09 |
|  | 6 months | 203 | 25.9 ± 15.5 | -4.1 (-5.7, -2.5) | **<0.001** |
|  | 18 months | * |  |  |  |
| AUDIT-C | Baseline | 264 | 2.66 ± 2.97 | 0 |  |
|  | 3 months | 173 | 2.48 ± 2.77 | 0.05 (-0.21, 0.31) | 0.70 |
|  | 6 months | 204 | 2.41 ± 2.62 | 0.03 (-0.21, 0.27) | 0.82 |
|  | 18 months | 199 | 2.27 ± 2.68 | -0.20 (-0.44, 0.04) | 0.10 |
| Variable | Timepoint | n | Mean score | Mean % change (from baseline) | P-value vs. baseline |
| Quality of Life  (EQ-5D-3L) | Baseline | 267 | 0.498 | - | - |
|  | 3 months | 167 | 0.527 | +5.88 | N/A |
|  | 6 months | 203 | 0.532 | +6.89 | N/A |
|  | 18 months | * |  |  |  |
| Variable | Timepoint | n | n (%) | Odds Ratio  (95% CI) | P-value vs. baseline |
| Drug use | Baseline | 264 | 24 (9.1%) | 1 |  |
|  | 3 months | 173 | 12 (6.9%) | 0.63 (0.20, 1.95) | 0.43 |
|  | 6 months | 204 | 20 (9.8%) | 1.59 (0.59, 4.24) | 0.36 |
|  | 18 months | 199 | 15 (7.5%) | 0.77 (0.27, 2.13) | 0.61 |

Table 1. Changes in key parameters over time. *Data not collected
