## Supplementary material for "Trajectory of severe COVID anxiety and predictors for recovery in an 18-month cohort": [Table 2 inserted here]

| COVID behaviour | Timepoint | n | n (%) | Odds ratio (95% CI) | P-value vs. baseline |
| --- | --- | --- | --- | --- | --- |
| Constant COVID worry | Baseline | 268 | 57 (21.3) | 1 | - |
|  | 3 months | 168 | 24 (14.3) | 0.62 (0.37-1.04) | 0.07 |
|  | 6 months | 203 | 26 (12.8) | 0.54 (0.33-0.90) | 0.02 |
|  | 18 months | 198 | 5 (2.5) | 0.10 (0.034-0.24) | <0.001 |
| Constantly watching COVID news | Baseline | 268 | 31 (11.6) | 1 | - |
|  | 3 months | 168 | 11 (6.5) | 0.54 (0.26-1.1) | 0.09 |
|  | 6 months | 203 | 9 (4.4) | 0.35 (0.16-.76) | 0.008 |
|  | 18 months | 198 | 4 (2.0) | 0.16 (0.054-0.45) | <0.001 |
| Never leaving home | Baseline | 268 | 31 (11.6) | 1 | - |
|  | 3 months | 168 | 14 (8.3) | 0.70 (0.36-1.35) | 0.28 |
|  | 6 months | 203 | 19 (9.4) | 0.79 (0.43-1.44) | 0.44 |
|  | 18 months | 198 | 10 (5.1) | 0.40 (0.19-0.85) | 0.02 |
| Not sending children to school* | Baseline | 94 | 19 (20.2) | 1 | - |
|  | 3 months | 53 | 10 (18.9) | 0.92 (0.39-2.15) | 0.84 |
|  | 6 months | 66 | 7 (10.6) | 0.46 (0.19-1.19) | 0.11 |
|  | 18 months | 61 | 7 (11.5) | 0.51 (0.20-1.30) | 0.16 |
| Buying all food online | Baseline | 268 | 100 (37.3) | 1 | - |
|  | 3 months | 168 | 61 (36.3) | 0.96 (0.64-1.43) | 0.83 |
|  | 6 months | 203 | 53 (26.1) | 0.59 (0.4-0.88) | 0.01 |
|  | 18 months | 198 | 39 (19.7) | 0.41 (0.27-0.63) | <0.001 |
| Constantly washing hands | Baseline | 268 | 54 (20.1) | 1 | - |
|  | 3 months | 168 | 41 (24.4) | 1.28 (0.81-2.03) | 0.30 |
|  | 6 months | 203 | 31 (15.3) | 0.71 (0.44-1.16) | 0.17 |
|  | 18 months | 198 | 23 (11.6) | 0.52 (0.31-0.88) | 0.02 |
| Washing all items coming into the home | Baseline | 268 | 83 (31.0) | 1 | - |
|  | 3 months | 168 | 39 (23.3) | 0.67 (0.43-1.04) | 0.08 |
|  | 6 months | 203 | 37 (18.2) | 0.50 (0.34-0.77) | 0.002 |
|  | 18 months | 198 | 24 (12.1) | 0.31 (0.19-0.51) | <0.001 |
| Washing each item of clothing when worn outside | Baseline | 268 | 47 (17.5) | 1 | - |
|  | 3 months | 168 | 29 (17.3) | 0.98 (0.59-1.63) | 0.94 |
|  | 6 months | 203 | 27 (13.3) | 0.72 (0.43-1.20) | 0.21 |
|  | 18 months | 198 | 21 (10.6) | 0.56 (0.32-0.97) | 0.04 |

Table 2. Changes in COVID-related thoughts and behaviours over time. *Of those respondents with school aged children.
