## Supplementary material for "Trajectory of severe COVID anxiety and predictors for recovery in an 18-month cohort": [Table 3 inserted here]

| **Factor / baseline score** | **Category** | **CAS Reduction** | **Univariable analysis** | | **Multivariable analysis** | |
| --- | --- | --- | --- | --- | --- | --- |
|  |  | **Mean ± SD** | **Coeff (95% CI)** | **P-value** | **Coeff (95% CI)** | **P-value** |
| Age (*) | - | - | -0.5 (-0.9, -0.2) | 0.005 | -0.4 (-0.7, -0.1) | 0.02 |
| Sex | Female | 8.4 ± 4.4 | 0 | 0.52 | - | - |
|  | Male | 9.7 ± 4.3 | -0.5 (-2.1, 1.0) |  |  |  |
| Ethnicity | Not at greater risk | 8.6 ± 4.3 | 0 | 0.06 | 0 | 0.002 |
|  | At greater risk | 9.0 ± 4.8 | -1.5 (-3.1, 0.9) |  | -2.2 (-3.6, -0.8) |  |
| Employment | Not employed | 8.4 ± 4.1 | 0 | 0.29 | - | - |
|  | Employed | 9.0 ± 4.6 | 0.6 (-0.5, 1.7) |  |  |  |
| Lives alone | No | 8.8 ± 4.3 | 0 | 0.07 | 0 | 0.08 |
|  | Yes | 7.8 ± 4.8 | -1.4 (-2.9, 0.1) |  | -1.2 (-3.6, -0.8) |  |
| Lives with vulnerable | No | 8.6 ± 4.5 | 0 | 0.23 | - | - |
| person | Yes | 8.8 ± 4.2 | -0.7 (-1.8, 0.4) |  |  |  |
| At risk condition | No | 8.7 ± 4.4 | 0 | 0.69 | - | - |
|  | Yes | 8.7 ± 4.1 | -0.3 (-1.5, 1.0) |  |  |  |
| Patient classification (§) | Cluster 1 – Simple | 8.0 ± 4.3 | 0 | 0.34 | - | - |
|  | Cluster 2 – At-risk | 8.8 ± 3.7 | 0.1 (-1.3, 1.4) |  |  |  |
|  | Cluster 3 - Anankastic | 8.5 ± 5.1 | -0.8 (-2.7, 1.0) |  |  |  |
|  | Cluster 4 - Complex | 10.1 ± 4.3 | 0.9 (-0.8, 2.5) |  |  |  |
| Vaccinated by 18 months | No  Yes | 8.6 ± 5.2  8.6 ± 4.3 | 0  1.0 (-0.7, 2.8) | 0.24 | - | - |
| CAS baseline | - | - | - |  | 0.7 (0.5, 0.9) | <0.001 |
| SAPAS baseline | < 4 | - | 0 | 0.77 | - | - |
|  | ≥ 4 | - | -0.2 (-1.3, 1.0) |  |  |  |
| GAD-7 baseline (^**^)  GAD-7 reduction (^**^) | -  - | -  - | 0.1 (-0.7, 0.9)  1.3 (0.8, 1.7) | 0.80  <0.001 | -  (***) | -  - |
| sHAI ^(*)^ baseline (^*^)  sHAI reduction (^*^) | -  - | -  - | -0.2 (-0.6, 0.2)  1.1 (0.7, 1.4) | 0.35  <0.001 | 0.8 (0.4, 1.2) | <0.001 |
| PHQ-9 baseline (^**^)  PHQ-9 reduction (^**^) | -  - | -  - | 0.2 (-0.4, 0.7)  1.1 (0.7, 1.4) | 0.54  <0.001 | -  0.8 (0.4, 1.2) | -  <0.001 |
| OCI-R baseline (^*^) | - | - | -0.2 (-0.6, 0.2) | 0.27 | - | - |
| AUDIT-C baseline | - | - | 0.0 (-0.2, 0.2) | 0.94 | - | - |

Table 3. Associations between demographic and clinical factors, and reduction in CAS scores from baseline to 18-month follow-up.

(§) Clusters were established with latent profile analysis, see King et al., 2023b.

(*) Regression coefficients reported for a 10-unit change in variable

(**) Regression coefficients reported for a 5-unit change in variable

(***) GAD-7 reductions were no longer associated with reducing CAS scores in multivariable analysis and so not included in the final model.

CAS COVID Anxiety Scale, *GAD-7* General Anxiety Disorder Assessment 7-item, *PHQ-9* Patient Health Questionnaire 9-item, *OCI-R* Obsessive-Compulsive Inventory Revised, *sHAI* Health Anxiety Inventory Short Form, *AUDIT-C* Alcohol Use Disorders Identification Test of Consumption, *SAPAS* Standardised Assessment of Personality Abbreviated Scale. Note: univariate analyses adjust for baseline CAS score.
